# Waves of COVID-19 pandemic. Detection and SIR simulations

**DOI:** 10.1101/2020.08.03.20167098

**Authors:** Igor Nesteruk

## Abstract

**Background:** Unfortunately, the COVID-19 pandemic is still far from stabilizing. Of particular concern is the sharp increase in the number of diseases in June-July 2020. The causes and consequences of this sharp increase in the number of cases are still waiting for their researchers, but there is already an urgent need to assess the possible duration of the pandemic, the expected number of patients and deaths. The resumption of international passenger traffic needs the information for deciding which countries’ citizens are welcome guests. Correct simulation of the infectious disease dynamics needs complicated mathematical models and many efforts for unknown parameters identification. Constant changes in the pandemic conditions (in particular, the peculiarities of quarantine and its violation, situations with testing and isolation of patients) cause various epidemic waves, lead to changes in the parameter values of the mathematical models.

**Objective:** In this article, pandemic waves in Ukraine and in the world will be detected, calculated and discussed. The estimations for hidden periods, epidemic durations and final numbers of cases will be presented. The probabilities of meeting a person spreading the infection and reproduction numbers will be calculated for different countries and regions.

**Methods:** We propose a simple method for the epidemic waves detection based on the differentiation of the smoothed number of cases. We use the known SIR (susceptible-infected-removed) model for the dynamics of the epidemic waves. The known exact solution of the SIR differential equations and statistical approach were modified and used.

**Results:** The optimal values of the SIR model parameters were identified for four waves of pandemic dynamics in Ukraine and five waves in the world. The number of cases and the number of patients spreading the infection versus time were calculated. In particular, the pandemic probably began in August 2019. If current trends continue, the end of the pandemic should be expected no earlier than in March 2021 both in Ukraine and in the world, the global number of cases will exceed 20 million. The probabilities of meeting a person spreading the infection and reproduction numbers were calculated for different countries and regions.

**Conclusions:** The SIR model and statistical approach to the parameter identification are helpful to make some reliable estimations of the epidemic waves. The number of persons spreading the infection versus time was calculated during all the epidemic waves. The obtained information will be useful to regulate the quarantine activities, to predict the medical and economic consequences of the pandemic and to decide which countries’ citizens are welcome guests.

## Introduction

Here we consider the global COVID-19 pandemic dynamics and epidemic outbreak in Ukraine with the use of official WHO data sets about the confirmed number of cases, [1]. The SIR model, connecting the number of susceptible *S*, infected and spreading the infection *I* and removed *R* persons, was applied in [2-4]. The unknown parameters of this model can be estimated with the use of the cumulative number of cases *V*=*I+R* and the statistics-based method of parameter identification [5, 6].

This approach was used in [6-16] to estimate the first waves of pandemic dynamics in Ukraine, Kyiv, China, the Republic of Korea, Italy, Austria, Spain, Germany, France, the Republic of Moldova, UK, USA and in the world. Usually the number of cases registered during the initial period of an epidemic is not reliable, since many infected persons are not detected. That is why the correct estimations of epidemic parameters can be done with the use of data sets obtained for later periods of the epidemic when the number of detected cases is closer to the real one. On the other hand, changes in quarantine conditions, human behavior, pathogen activity, weather etc. can cause changes in the course of the pandemic, namely the so-called epidemic waves. The mathematical simulation of these waves needs development some criteria of the waves detection, modification of models and methods of parameter identification.

In this paper we will update the estimations of the first waves of the pandemic dynamics in Ukraine and in the world with the use of SIR model [2-4] and the statistics-based method of parameter identification [5, 6]. Simple criteria for identification of next pandemic waves will be proposed. The SIR model and the parameter identification procedure will be modified in order to simulate next waves of the pandemic. The results of calculations for 5 global waves of Covid-19 pandemic and 4 waves of the epidemic in Ukraine will be presented and discussed. The new estimations for probabilities of meeting a person spreading the infection and for reproduction numbers will be proposed.

## Materials and Methods

### Data

The official information regarding the accumulated numbers of confirmed COVID-19 cases *V*_*j*_ in Ukraine and in the world from WHO daily situation reports (numbers 101-194), [1] is presented in Table 1. The corresponding moments of time *t*_*j*_ (measured in days, zero point corresponds to January 20, 2020) are also shown in this table. Some time periods were used for calculations. Other values were used only for verifications of their results.

**Table 1.**
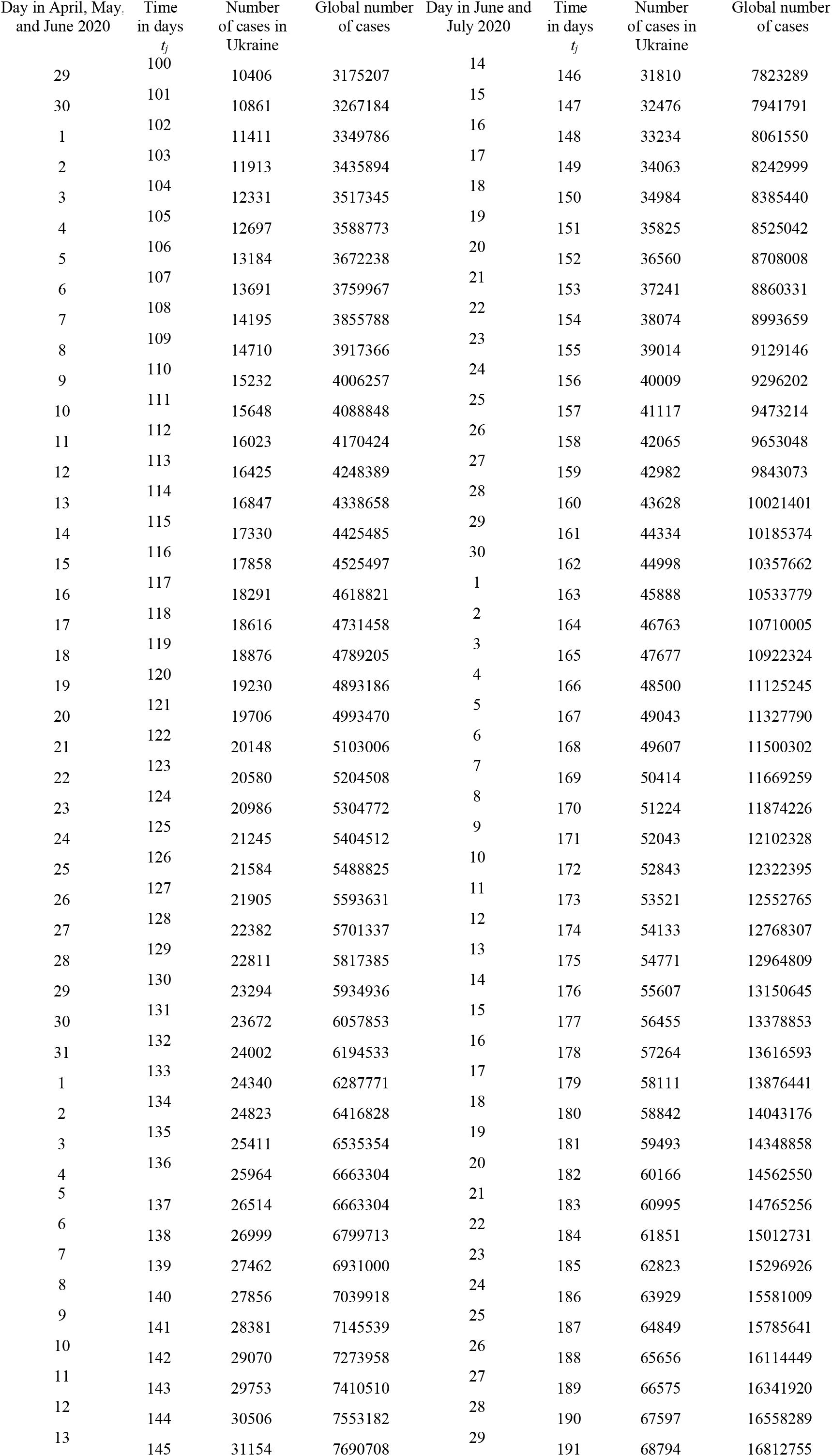
Official cumulative numbers of confirmed cases in Ukraine and in the world, [1].

### Epidemic waves detection

Changes in quarantine conditions, human behavior, pathogen activity, weather etc. can cause changes in the epidemic dynamics, namely the so-called epidemic waves. The simplest way of their detection is to find some changes in the dependences of the number of registered cases on time. Since the number of cases is random, its time dependence needs some smoothing. We can use the method proposed in [16]:

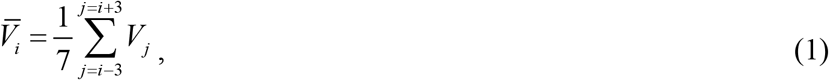

and the derivatives of the smoothed values

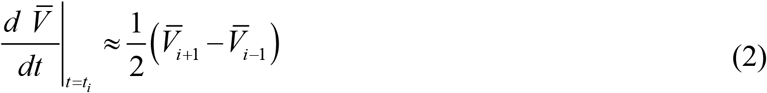

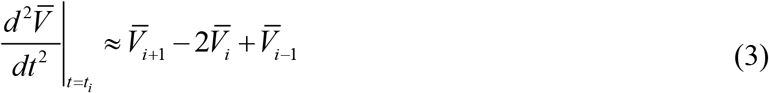

in order to detect the changes in epidemics dynamics.

### SIR model

The SIR model for an infectious disease [2-5] relates the number of susceptible persons *S* (persons who are sensitive to the pathogen and **not protected**); the number of infected is *I* (persons who are sick and **spread the infection**; please don’t confuse with the number of still ill persons, so known active cases) and the number of removed *R* (persons who **no longer spread the infection**; this number is the sum of isolated, recovered, dead, and infected people who left the region):

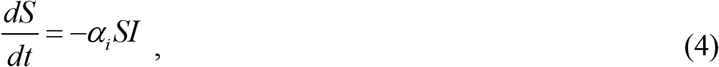

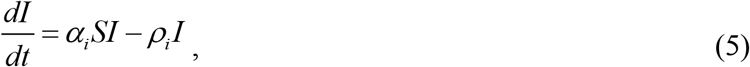

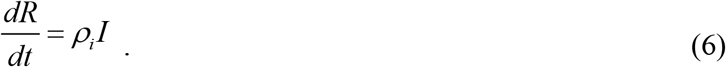

Parameters *α*_*i*_ and *ρ*_*i*_ are supposed to be constant for every wave of the epidemics, i.e. for the time periods *t*_*i*_ ≤ *t* ≤ *t*_*i*+1_, *i* = 1, 2,3,….

Parameters *α*_*i*_ show how quick the susceptible persons become infected (see (4)). Large values of this parameter correspond to severe epidemics with many victims. These parameters accumulate many characteristics. First they shows how strong (virulent) is the pathogen and what is the way of its spreading. Parameters *α*_*i*_ accumulates also the frequency of contacts and the way of contacting. In order to decrease the values of *α*_*i*_, we have to minimize the number of our contacts and change our contacting habits. For example, we have to avoid the public places and use masks there, minimize or cancel traveling. We have to change our contact habits: to avoid handshakes and kisses. First, all these simple things are very useful to protect yourself. In addition, if most people follow these recommendations, we have chance to diminish the values of parameter *α*_*i*_ and reduce the negative effects of the pandemic.

The parameters *ρ*_*i*_ characterize the patient removal rates, since eq. (6) demonstrates the increase rate for *R*. The inverse values 1/ *ρ*_*i*_ are the estimations for time of spreading the infection *τ*. So, we are interested in increasing the values of parameters *ρ*_*i*_ and decreasing 1/ *ρ*_*i*_. People and public authorities should work on this and organize immediate isolation of suspicious cases.

Since the derivative *d* (*S* + *I* + *R*) / *dt* is equal to zero (it follows from summarizing Eqs. (4)-(6)), the sum

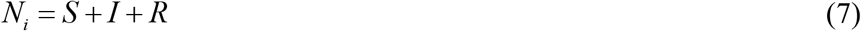

must be constant for every wave and is not the volume of population (see also [15]).

### Analytical solution of SIR equations

To determine the initial conditions for the set of equations (4)–(6), let us suppose that at the beginning of every epidemic wave *t*_*i*_ :

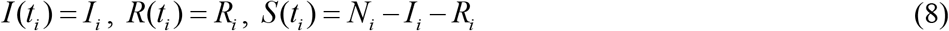

In particular, when the first wave of the epidemic starts with one infected person, the initial conditions (8) can be written as follows:

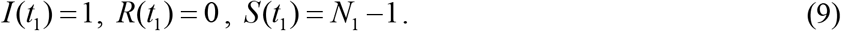

Equations (9) were used in [5-16] to simulate the first waves of epidemics in different countries.

It follows from (4) and (5) that

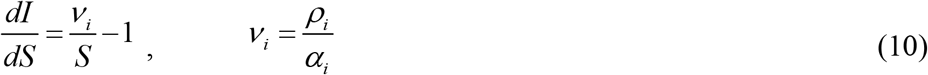

Integration of (10) with the initial conditions (8) yields:

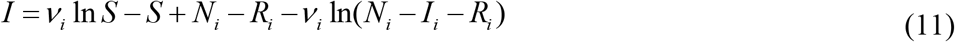

It follows from (11) that function *I* has a maximum at *S* =*ν*_*i*_ and tends to zero at infinity. The corresponding number of susceptible persons at infinity *S*_*i*∞_ > 0 can be calculated from a non-linear equation

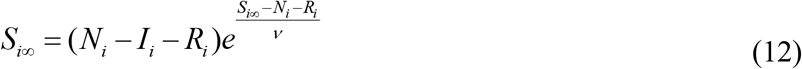

Formula (12) follows from (11) at *I*=0.

As in [5] we solve (4)-(6) by introducing the function *V* (*t*) = *I* (*t*) + *R*(*t*), corresponding to the number of victims or cumulative confirmed number of cases. It follows from (5)-(7) and (11) that:

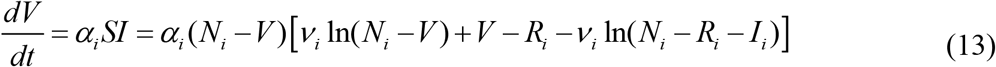

Integration of (13) yield an analytical solution for the set of Equations (4)–(6):

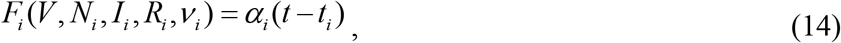

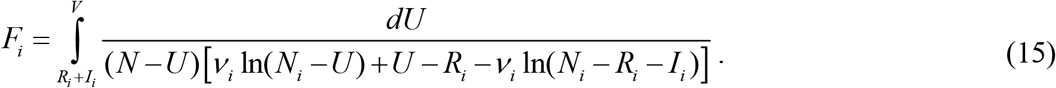

Thus, for every set of parameters *N*_*i*_, *I*_*i*_, *R*_*i*_, *ν*_*i*_, *α*_*i*_, *t*_*i*_ and a fixed value of *V*, integral (15) can be calculated and a corresponding moment of time can be determined from (14). Then functions *I*(*t*) and *R*(*t*) can be easily calculated with the of formulas (11) and:

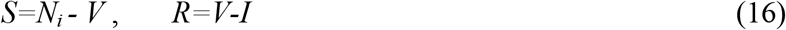

The final number of victims (final accumulated number of cases) can be calculated from:

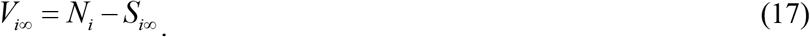

To estimate the duration of an epidemic outbreak, we can use the condition:

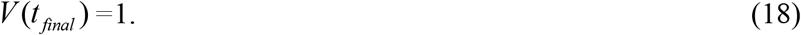

which means that at *t* > *t* _*final*_ less than one person still spreads the infection.

### Parameter identification procedure

In the case of a new epidemic, the values of its independent six parameters are unknown and must be identified with the use of limited data sets. For the first wave of an epidemic starting with one infected person, the number of unknown parameters is only four, since *I*_1_ = 1 and *R*_1_ = 0. The corresponding statistical approach was used in [6-16] to estimate the values of four unknown parameters for the first waves of Covid-19 pandemic in different regions.

For the next epidemic waves (*i* > 1), the registered number of victims *V*_*j*_ corresponding to the moments of time *t*_*j*_ can be used in eq. (15) in order to calculate *F*_*i, j*_ = *F*_*i*_ (*V*_*j*_, *N*_*i*_, *ν*_*i*_, *I*_*i*_, *R*_*i*_) for every fixed values of *N*_*i*_, *ν* _*i*_, *I*_*i*_, *R*_*i*_ and then to check how the registered points fit the straight line (14). For this purpose the linear regression can be applied, e.g., [17], and the optimal straight line, minimizing the sum of squared distances between registered and theoretical points, can be defined. Thus we can find the optimal values*α*_*i*_, *t*_*i*_ and calculate the correlation coefficient *r*_*i*_ for the of linear dependence (14).

Then the F-test may be applied to check how the null hypothesis that says that the proposed linear relationship (14) fits the data set. The experimental values of the Fisher function can be calculated with the use of the formula:

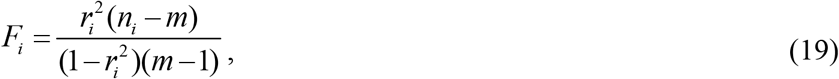

where *n*_*i*_ is the number of observations for the *i*-th wave of epidemis, *m* = 2 is the number of parameters in the regression equation, [17]. The corresponding experimental value *F*_*i*_ has to be compared with the critical value *F*_*C*_ (*k*_1_, *k*_2_) of the Fisher function at a desired significance or confidence level (*k*_1_ = *m* −1, *k*_2_ = *n*_*i*_ − *m*), [18]. When the values *n*_*i*_ and *m* are fixed, the maximum of the Fisher function coincides with the maximum of the correlation coefficient. Therefore, to find the optimal values of parameters *N*_*i*_, *ν*_*i*_, *I*_*i*_, *R*_*i*_, we have to find the maximum of the correlation coefficient for the linear dependence (14). To compare the reliability of different predictions (with different values of *n*_*i*_) it is useful to use the ratio *F*_*i*_ / *F*_*C*_ (1, *n*_*i*_ − 2) at fixed significance level. We will use the level 0.001; corresponding values of *F*_*C*_ (1, *n*_*i*_ − 2) can be taken from [18]. The most reliable prediction yields the highest *F*_*i*_ / *F*_*C*_ (1, *n*_*i*_ − 2) ratio.

In the case of sequential calculation of epidemic waves *i* = 1,2,3 …, it is possible to avoid determining the four optimal unknown parameters *N*_*i*_, *ν*_*i*_, *I*_*i*_, *R*_*i*_, thereby reducing the amount of calculations. For parameters *I*_*i*_, *R*_*i*_ it is possible to use the numbers of *I* and *R* calculated for the previous wave of epidemic at the moment of time when the following wave began. In this study we will use this approximate approach.

## Results

### Detection of Covid-19 pandemic waves in Ukraine and in the world

Applications of formulae (1)-(3) for the pandemic dynamics in Ukraine and in the world are shown in Fig. 1 and 2. The accumulated numbers of cases (blue “circles”) were smoothed with the use of eq. (1) and shown by blue lines. Red and black markers represent the results of differentiation (2) and (3) respectively. The make the results more visible, the first derivative (2) is multiplied by 100, the second one (formula (3)) – by 1000.

**Fig 1.**
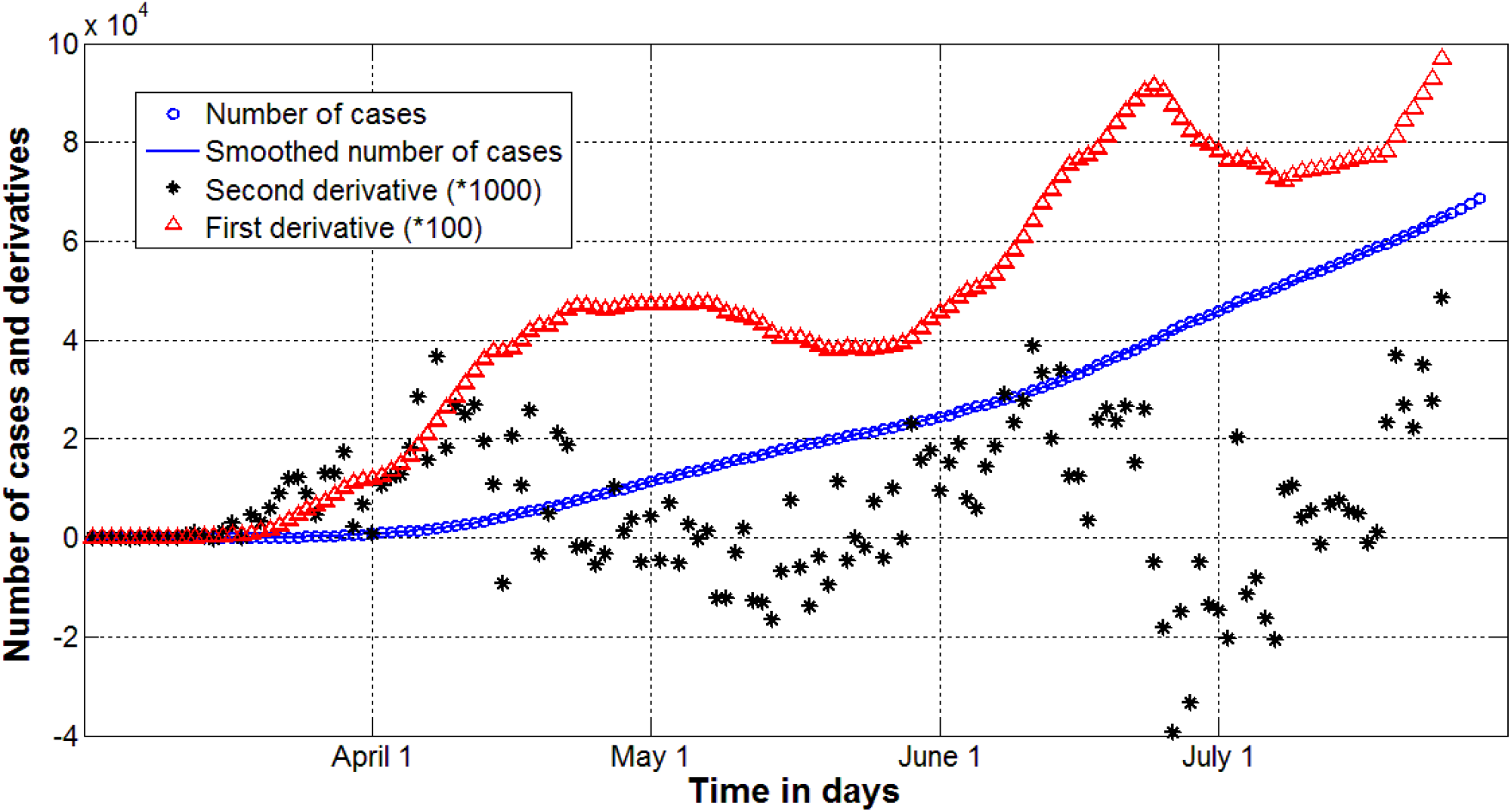
Epidemic dynamics in Ukraine versus time in days. Accumulated number of cases (blue markers and line, eq. (1)). Red markers represent first derivative (eq. (2)), black one show the second derivative (eq. (3)).

**Fig 2.**
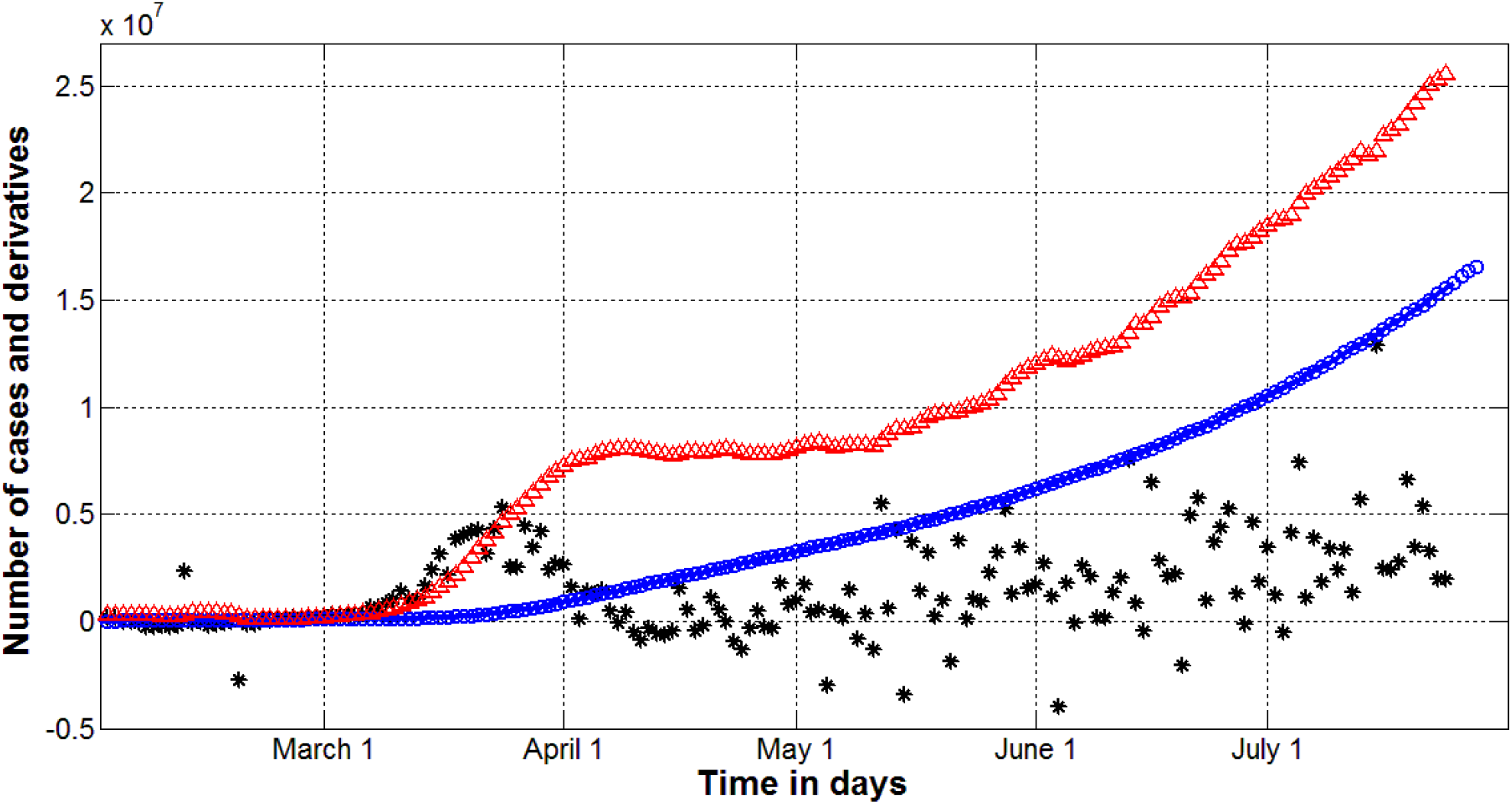
Pandemic dynamics in the world versus time in days. Accumulated number of cases (blue markers and line, eq. (1)). Red markers show first derivative (eq. (2)) multiplied by 100, black one - the second derivative (eq. (3)) multiplied by 1000.

Fig. 1 and 2 demonstrate that the second derivatives increase after epidemic outbreak, then become smaller and negative. Such behavior is typical for the first wave of the epidemics (before May 17, 2020 in Ukraine, see Fig. 1 and before May 12 in the world, see Fig. 2). The jumps in the values of the second derivative indicate changes in the conditions of the pandemic (for example, the weakening of quarantine) and the transitions to the next waves with other values of the parameters of mathematical models. These jumps occurred on May 16, May 29, June 8, July 3 and July 19-20 in Ukraine (see Fig. 1) and on May 12 and 28; June 13; July 5 and 14 in the world (see Fig. 2). Therefore these days can be treated as the beginning of the second, third etc. waves.

Thus four waves were calculated for Ukraine and presented in the next Section. The distance between some jumps of the second derivative, which took place in the second half of May, was too small to make statistical estimates of the parameters, so the individual waves of the epidemic during this period were not isolated. The same situation occurred for the world dynamics in July. The distance between jumps of second derivative was too short in order to isolate two different waves. Thus five waves were calculated for the world pandemic dynamics. These facts can reduce the accuracy of SIR simulations. At the end of June 2020, the number of days of observation of the fifth wave in Ukraine (after July 19) was still insufficient for statistical analysis, so this wave can be calculated later.

The second epidemic wave in Ukraine was caused by the cancelation of the national lockdown on May 10. After the incubation period (approximately after May 16), the number of new cases began to grow faster. Further waves of the epidemic in Ukraine are associated with further easing of quarantine and mass non-compliance with social distancing. The fifth epidemic wave in Ukraine can be explained by the consequences of the holiday season, which increased the number of trips and violations of social distancing. Analyze the causes of new waves in the world is difficult because of the large number of countries with different pandemic dynamics. Probably the fourth pandemic wave after June 13 is associated with mass protests in the United States.

## Results of SIR simulations

The first waves of the pandemic in Ukraine and in the world were already simulated with the use SIR model [11-16]. Usually the number of cases during the initial period of a new epidemic outbreak is not reliable, since many cases are not detected. That is why the first waves need re-estimations after obtaining fresh data. In this paper we have recalculated the first waves of pandemic in Ukraine and in the world with the use of datasets for the periods of time immediately preceding the second wave. The characteristics of waves 2-4 for Ukraine and pandemic waves 2-5 in the world were also calculated and presented in Tables 2-5. Value *n*_*i*_ =14 was used for all the cases.

**Table 2.** Characteristics of the first epidemic wave in Ukraine calculated with the use of different data sets. Optimal values of parameters, final sizes and days (last three rows).

| **Characteristics** | **Ukraine,**<br>**first wave,**<br>**prediction 8, [13]**<br>**i=1** | **Ukraine,**<br>**first wave,**<br>**prediction 19, [16]**<br>**i=1** | **Ukraine,**<br>**first wave,**<br>**prediction 21**<br>**i=1** |
| --- | --- | --- | --- |
| Period taken for calculations | April 11-24 | May 13-26 | May 3-16 |
| $I_i$ | 1 | 1 | 1 |
| $R_i$ | 0 | 0 | 0 |
| $N_i$ | 21380.9241600000 | 92770.3987200000 | 59002.368 |
| $\nu_i$ | 12190.2413389748 | 76647.6354922344 | 43307.0168670536 |
| $\alpha_i$ | 1.45792603472e-05 | 3.766268949867e-06 | 4.62451395337e-06 |
| $t_i$ | 27.72672361552338 | -32.4672286400970 | -16.5560950369874 |
| $\rho_i$ | 0.177724702176649 | 0.288675609635095 | 0.200273903780639 |
| $1/\rho_i$ | 5.62667984671 | 3.46409591466375 | 4.99316177056849 |
| $r_i$ | 0.998863416866565 | 0.998856483326519 | 0.999453409358706 |
| $F_i$, eq. (19) | 5269.98203160936 | 5237.97374486281 | 10968.1371753815 |
| $F_i / F_C(1, n_i - 2)$ | 283.332367290826 | 281.611491659291 | 589.684794375351 |
| $S_{i\infty}$, eq. (12) | 6108 | 62507 | 30676 |
| $V_{i\infty}$, eq. (17) | 15273 | 30263 | 28327 |
| $t_{final}$, eq. (18) | 186.5 | 273.6 | 264.9 |
| Final day of epidemic | July 26, 2020 | October 20, 2020 | October 11, 2020 |

To illustrate the influence of data on the results of SIR simulations, the previous estimations of the first waves are also presented in Tables 2 and 4. It can be seen, that the use of more recent (and complete) data has changed the estimation for the pandemic beginning. Table 2 illustrates that prediction 21 calculated with the use of number of cases from the period May 3-16 (immediately before the start of the second wave) yields much longer hidden period of the epidemic outbreak in Ukraine in comparison with the previous prediction 8, [13]. Prediction 19, [16] yields even longer hidden period, but it was obtained with the use of the dataset for the period May 13-26, which corresponds the transition form first to the second wave. This prediction yielded the smallest values of *F*_*i*_ and *F*_*i*_ / *F*_*C*_ (1, *n*_*i*_ − 2) in comparison with predictions 8 and 21 (see Table 2). The maximum corresponding values of these parameters demonstrate that the prediction 21 estimating the epidemic outbreak in Ukraine in the beginning of January, 2020 is probably the most reliable.

**Table 3.** Characteristics of second, third and fourth epidemic waves in Ukraine. Optimal values of parameters, final sizes and days (last three rows).

| Characteristics | Ukraine,<br>second wave,<br>i=2 | Ukraine,<br>third wave,<br>i=3 | Ukraine,<br>fourth wave,<br>i=4 |
| --- | --- | --- | --- |
| Period taken for calculations | May 17-30 | June 9-22 | July 3-16 |
| $I_i$ | 2196 | 2490 | 3736 |
| $R_i$ | 16420 | 25891 | 43941 |
| $N_i$ | 239587.2 | 149444.16 | 182294.495539200 |
| $\nu_i$ | 209382.9208704 | 104004.527376384 | 119723.766724183 |
| $\alpha_i$ | 7.728220925e-07 | 2.117564829222e-06 | 1.35741149806184e-06 |
| $t_i$ | 118.126430079990 | 140.948538546930 | 164.986258791213 |
| $\rho_i$ | 0.161815747046830 | 0.220236329252125 | 0.162514417542680 |
| $1/\rho_i$ | 6.17986826529682 | 4.54057694929708 | 6.15330021250195 |
| $r_i$ | 0.998894780369877 | 0.999714332158213 | 0.999456710370030 |
| $F_i$, eq. (19) | 5419.78688520125 | 20994.4146122108 | 11034.8339181942 |
| $F_i / F_C(1, n_i - 2)$ | 291.386391677487 | 1128.73196839843 | 593.270640763128 |
| $S_{i\infty}$, eq. (12) | 178641 | 78509 | 89552 |
| $V_{i\infty}$, eq. (17) | 60946 | 70935 | 92742 |
| $t_{final}$, eq. (18) | 531.9 | 338.5 | 421.7 |
| Final day of epidemic | July 5, 2021 | December 24, 2020 | March 17, 2021 |

**Table 4.** Characteristics of the first and second pandemic waves in the world. Two datasets for different periods of time were used to simulate the first wave. Optimal values of parameters, final sizes and days (last three rows).

| Characteristics | World,<br>first wave,<br>prediction 1, [15]<br>$i=1$ | World,<br>first wave,<br>prediction 2<br>$i=1$ | World,<br>second wave,<br>$i=2$ |
| --- | --- | --- | --- |
| Period taken for calculations | April 9-29 | April 29 - May 12 | May 12-25 |
| $I_i$ | 1 | 1 | 295912 |
| $R_i$ | 0 | 0 | 3874512 |
| $N_i$ | 6637317.12 | 23600000 | 113600000 |
| $\nu_i$ | 3537150.79869746 | 19925870.4384000 | 102947264.512000 |
| $\alpha_i$ | 2.34901375364e-08 | 1.380196708625e-08 | 2.66429099926749e-09 |
| $t_i$ | -111.265486657790 | -170.837434239689 | 113.050342136891 |
| $\rho_i$ | 0.0830881587484244 | 0.275016207955738 | 0.274281470238531 |
| $1/\rho_i$ | 12.0354093178044 | 3.63614932891861 | 3.64588974650873 |
| $r_i$ | 0.999861835424859 | 0.999848053500794 | 0.999760778228936 |
| $F_i$, eq. (19) | 68744.3303788117 | 39478.5832347979 | 25072.3295928952 |
| $F_i / F_C(1, n_i - 2)$ | 4522.65331439551 | 2122.50447498914 | 1347.97470929544 |
| $S_{i\infty}$, eq. (12) | 1595881 | 16654425 | 93209515 |
| $V_{i\infty}$, eq. (17) | 5041436 | 6945575 | 20390485 |
| $t_{final}$, eq. (18) | 393 | 415.5 | 749.5 |
| Final day of epidemic | February 16, 2021 | March 11, 2021 | February 18, 2022 |

The transitional period from first to second wave was used in [8] to estimate the characteristics of the epidemic in the Republic of Korea. In the next calculation for this country [14, 15] the dataset corresponding the second wave was used, but the first wave approach (*I*_*i*_ =1, *R*_*i*_=0) was applied. These facts reduce the accuracy of both estimates for South Korea.

Table 4 illustrates that the use of more recent (and complete) data changed the estimation for the pandemic beginning in the world. Probably, it happened in China in the beginning of August 2019. It must be taken into account that dataset for the period April 29 - May 12 was taken for calculations. On the one hand, this dataset most fully reflects the number of identified cases, but on the other hand, during the period from August 2019 to May 2020, the conditions of the pandemic could change, so such estimates of the date of the pandemic beginning should be treated critically.

The results of SIR simulations of the next waves of the pandemic in Ukraine and in the world are shown in Tables 3-5 and in Figs. 3 and 4. In can be seen that optimal values of the model parameters are rather different for different pandemic waves. In particular, the final sizes and durations of the pandemic significantly differ. It is not surprising, since different time periods with different conditions were used for calculations. The optimal calculated values of *t*_*i*_ are very close to the moments of time, when the corresponding wave has started.

**Fig 3.**
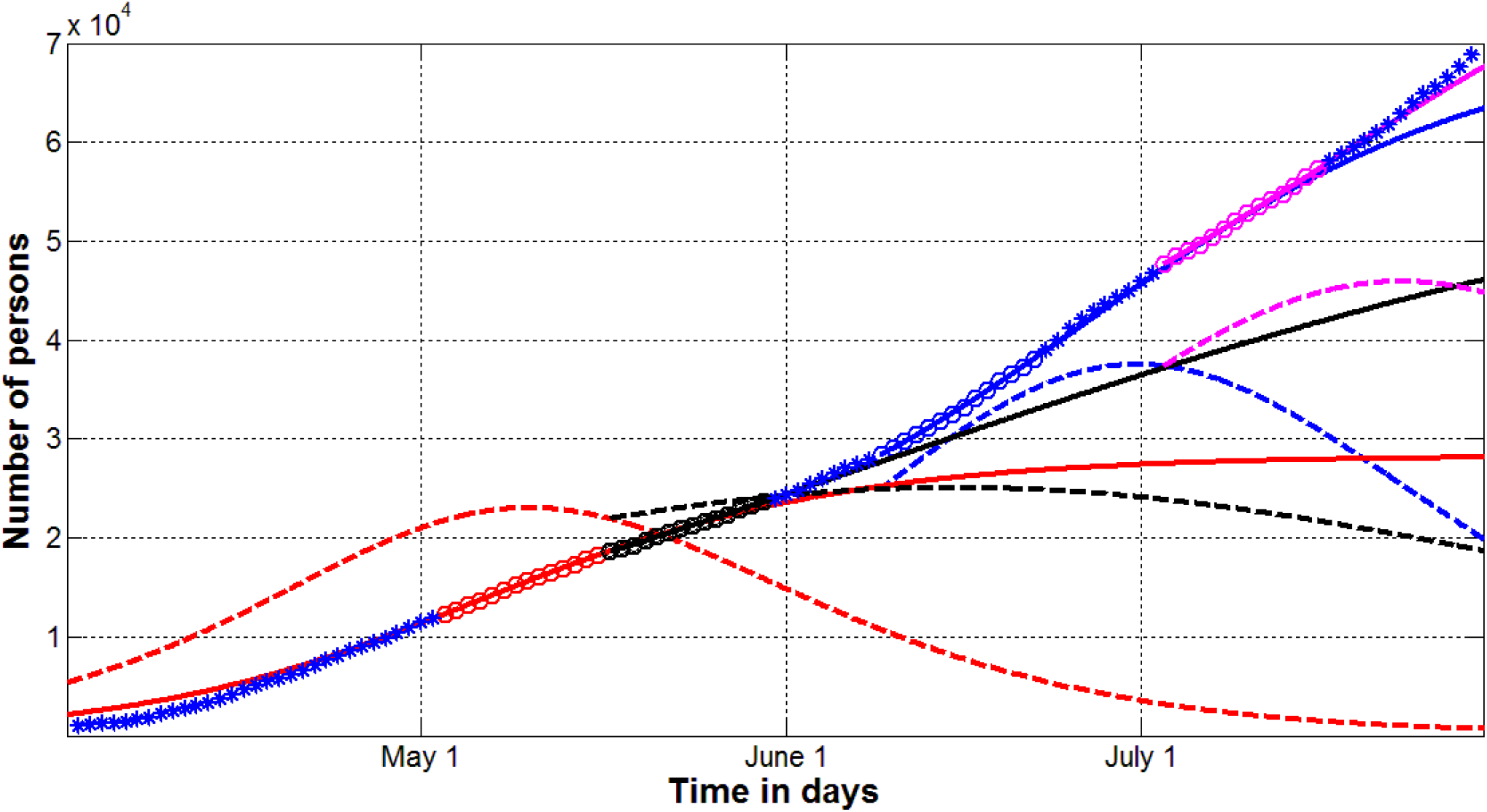
Epidemic waves in Ukraine. SIR curves (lines) and accumulated number of cases (markers) versus time. Red, black, blue and magenta colors correspond to waves 1, 2, 3 and 4 respectively. Numbers of infected and spreading *I*10* (dashed lines) and victims *V*=*I+R* (solid lines). “Circles” represent the values *V*_*j*_ taken for calculations, “stars” show the accuracy of calculations.

**Fig 4.**
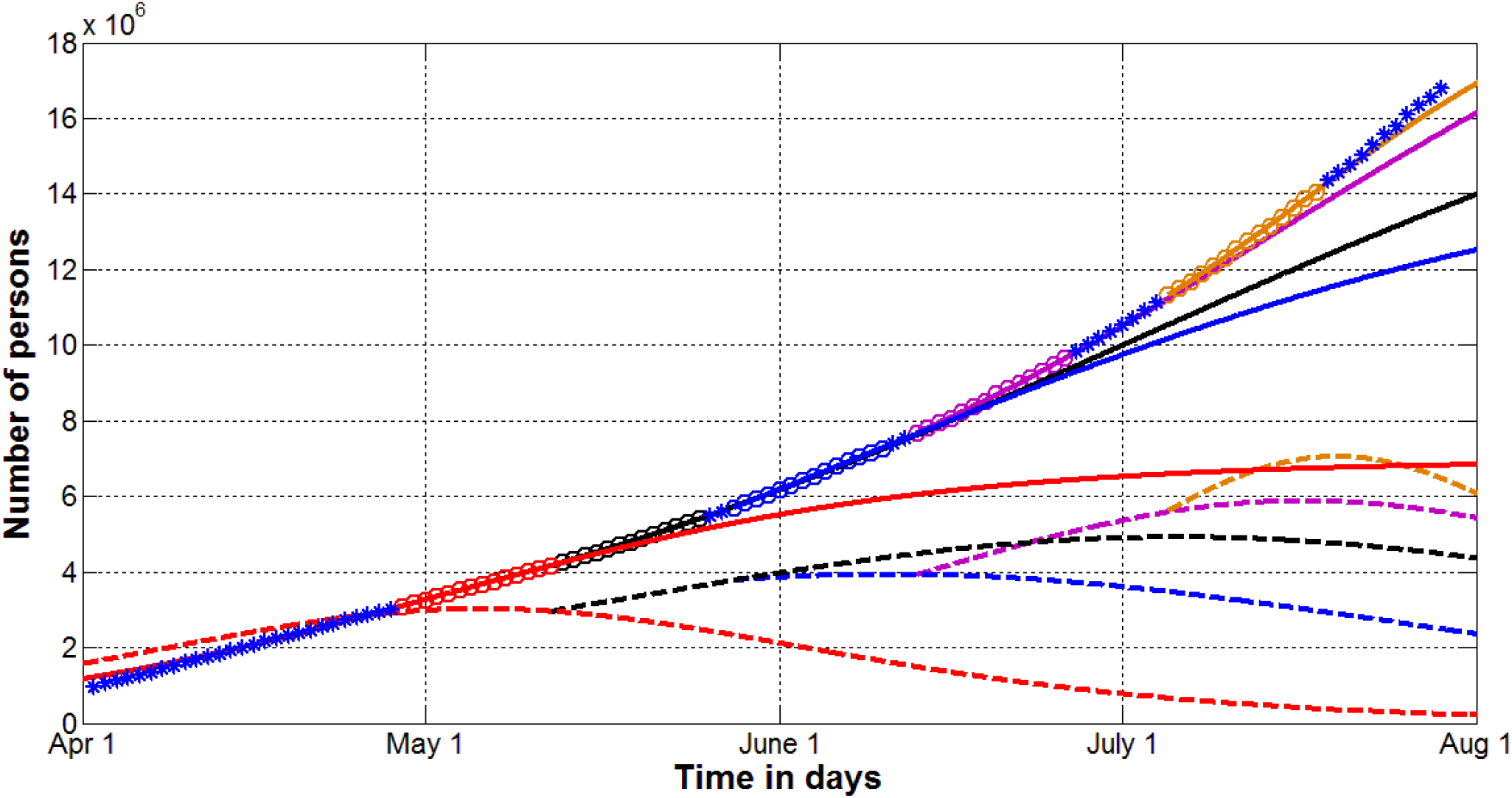
Pandemic waves in the world. SIR curves (lines) and accumulated number of cases (markers) versus time. Red, black, blue, magenta and brown colors correspond to waves 1, 2, 3, 4 and 5 respectively. Numbers of infected and spreading *I*10* (dashed lines) and victims *V*=*I+R* (solid lines). “Circles” represent the values *V*_*j*_ taken for calculations, “stars” show the accuracy of calculations.

Figs. 3 and 4 show four waves of the epidemic in Ukraine and five pandemic waves in the world. The accumulated number of cases *V*=*I+R* (solid lines) increases for each next wave. Every new wave also increases the number of infected and spreading the infection persons *I*. The calculated dependences *10*I*(*t*) are shown by dashed lines. The accuracy of simulations is rather good, especially for second and further waves of pandemic, since blue “stars” (showing the accumulated values of confirmed cases used only for control the calculations) are located very close to the corresponding solid lines showing the calculated *V*=*I+R* values. There are some discrepancies for the early stages of first waves, when the number of registered cases is lower that the real one due to the problems with the identification of the infected persons. It can be seen also the beginning of the fifth wave in Ukraine. After July 23, the real number of cases is higher than the calculated one for the fourth wave (compare blue “stars” and solid magenta line in Fig. 3).

The quarantine releasing in Ukraine caused new waves of the epidemic and the increase of the infected persons number. Unfortunately, this number is still around its maximal value in Ukraine (see magenta dashed line in Fig. 3). The fifth epidemic wave (which has to be calculated later) will probably show further increase in the number of persons spreading the infection.

## Discussion

### Hidden periods of Covid-19 pandemic

It was already mentioned that during the initial stages of epidemic the registered number of cases is much lower than the real one, since there are a lot of infected persons without symptoms. In every country some hidden periods occurred, before the first case was confirmed [14, 15]. To make reliable estimations of hidden periods, we have to use the datasets corresponding to the period when the number of registered cases approaches the real one during the first wave of epidemic. In prediction 21 for Ukraine (see Table 2) and in prediction 2 for the world (see Table 4), the periods of time just before the start of the second wave were used for SIR calculations. The results are shown in Fig. 5.

**Fig 5.**
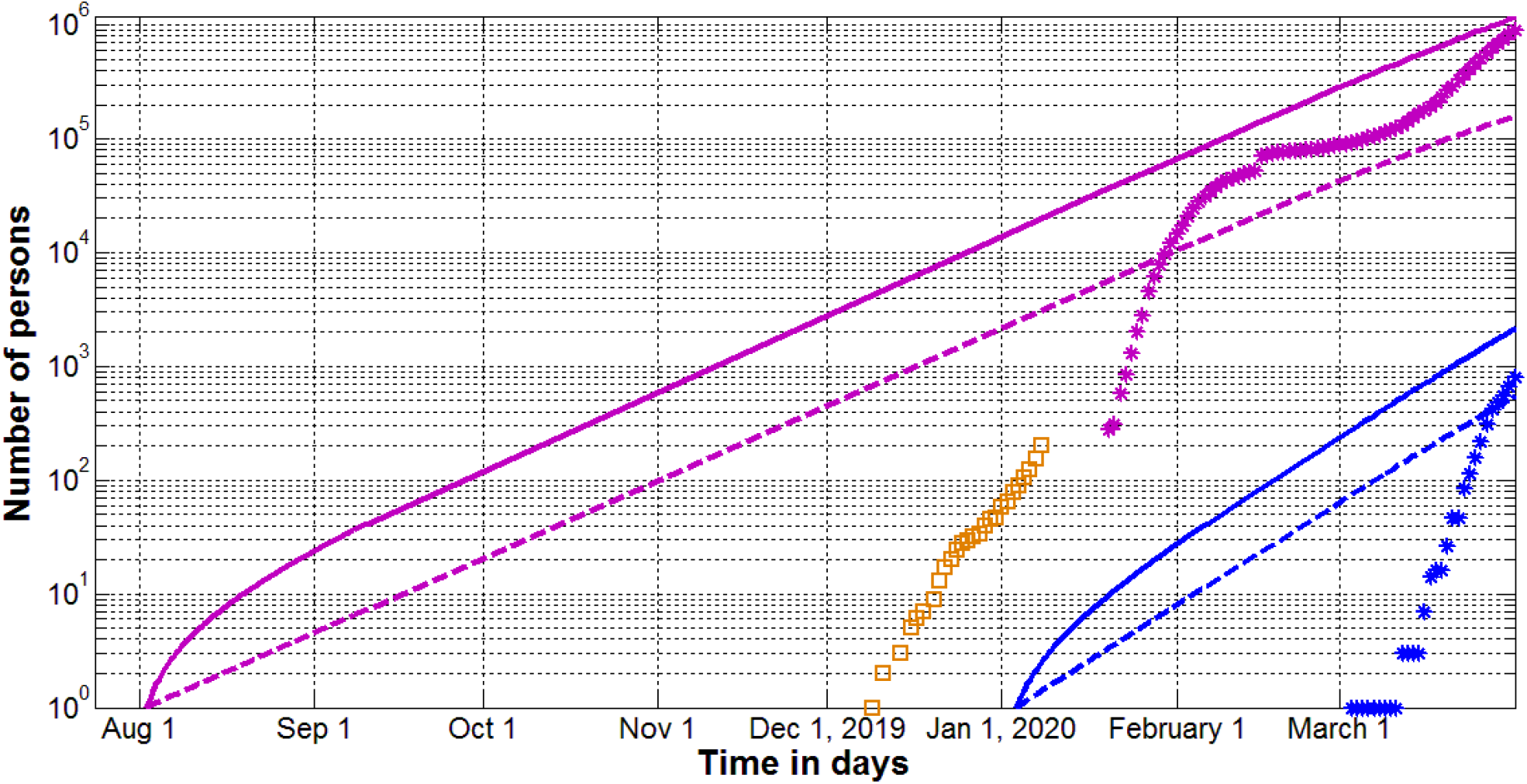
Hidden periods in Ukraine and in the world. SIR curves for the first wave of pandemic (lines) and accumulated number of cases (markers) versus time. Blue and magenta colors correspond to Ukraine (Table 1) and the world (Table 2) respectively. Numbers of infected and spreading *I* (dashed lines) and victims *V*=*I+R* (solid lines). “Stars” represent the accumulated number of cases *V*_*j*_ from [1]; “squares” show the number of laboratory confirmed cases in Wuhan, China calculated in [9] with the use of daily data presented in [19].

In particular the hidden period in the world according to the second simulation of the first wave is approximately 60 days longer in comparison this the first prediction for the first wave (see Table 4, [15]). Ukrainian statistics yields similar results. Estimation No. 8 available in [13-15] yielded February 16 as the day of the epidemic outbreak in Ukraine. The results of calculations, performed with the use of the more recent dataset (days immediately preceding the beginning of the second wave) and shown in Table 2, demonstrate that the first cases in Ukraine probably appeared in the beginning of January, 2020 (see solid blue line in Fig. 5).

Solid magenta line in Fig. 5 demonstrates that Covid-19 probably started to spread in August 2019. On December 31, 2019 – the day when China notified WHO about the situation in Wuhan - more than 2,000 persons could spread the infection (see dashed magenta line in Fig. 5). In the period from 18 to 27 October, 2019 (when the Military World Games held in Wuhan with the participation of 9,300 athletes from more than 140 countries) the number of infected persons can be estimated by 50-80 (see dashed magenta line in Fig. 5). May be some participants got the infection and passed it on to their families, [20].

### Long-term predictions for Ukraine and the world

Table 6 illustrates a rather high accuracy of calculations. It can be seen that after one week of observations (after the last day of the periods taken for calculations) the real accumulated number of cases exceed the calculated ones by 0.54% and 1.35% for Ukraine and the world respectively. After two weeks of observations the discrepancies are higher (3.31% for Ukraine and 4.25% for the world).

**Table 5.** Characteristics of the third, fourth and fifth pandemic waves in the world. Optimal values of parameters, final sizes and days (last three rows).

| Characteristics | World,<br>third wave,<br>i=3 | World,<br>fourth wave,<br>i=4 | World,<br>fifth wave,<br>i=5 |
| --- | --- | --- | --- |
| Period taken for calculations | May 28 – June 10 | June 13-26 | July 5-18 |
| $I_i$ | 377816 | 392933 | 562468 |
| $R_i$ | 5323521 | 7297775 | 10765332 |
| $N_i$ | 95388160 | 94880000 | 47113472 |
| $\nu_i$ | 88021224.0422502 | 81424517.3514240 | 32637843.7070307 |
| $\alpha_i$ | 3.5269848715e-09 | 3.903409800432e-09 | 9.51859566656283e-09 |
| $t_i$ | 129.027798384899 | 145.076908463997 | 167.113076654722 |
| $\rho_i$ | 0.310449525572027 | 0.317833259025022 | 0.310666437675697 |
| $1/\rho_i$ | 3.22113553936803 | 3.14630382945943 | 3.21888649279808 |
| $r_i$ | 0.999820043845012 | 0.999654631292158 | 0.999700105970476 |
| $F_i$, eq. (19) | 33332.4550282053 | 17363.7383017448 | 19998.0676446882 |
| $F_i / F_C(1, n_i - 2)$ | 1792.06747463469 | 933.534317298105 | 1075.16492713378 |
| $S_{i\infty}$, eq. (12) | 79959644 | 72028177 | 26315036 |
| $V_{i\infty}$, eq. (17) | 15428516 | 22851823 | 20798436 |
| $t_{final}$, eq. (18) | 652.3 | 600 | 427.2 |
| Final day of epidemic | November 2, 2021 | September 11, 2021 | March 22, 2021 |

**Table 6.** Comparison of the theoretical results with the real number of cases after one and two weeks of observations.

| Characteristics | Ukraine,<br>fourth wave,<br>July 23, 2020 | Ukraine,<br>fourth wave,<br>July 30, 2020 | World,<br>fifth wave,<br>July 25, 2020 | World,<br>fifth wave,<br>August 1, 2020 |
| --- | --- | --- | --- | --- |
| Calculated number of cases $V=I+R$ | 62486 | 67571 | 15571846 | 16909569 |
| Confirmed number of cases, WHO reports, [1] | 62823 | 69884 | 15785641 | 17660523 |
| Discrepancy (%) | 0.54 | 3.31 | 1.35 | 4.25 |

Fig. 6 shows the long-term predictions for Ukraine and the world for the period August-December, 2020. Despite the relatively high accuracy of pandemic wave calculations (see Table 6), these long-term forecasts given should be considered as preliminary and overly optimistic. The new waves are expected due to the quarantine easing and changes in social distancing. Gradual weakening of quarantines may not cause pronounced new waves. In this case, the parameters of the pandemic dynamics will change continuously, which makes the proposed approach unsuitable. The expected emergence of vaccines could also change the course of the pandemic. If mass vaccination begins in 2021, the total number of cases will still exceed 20 million. In any case the global and Ukrainian dynamics must be updated with the use of new data sets.

**Fig 6.**
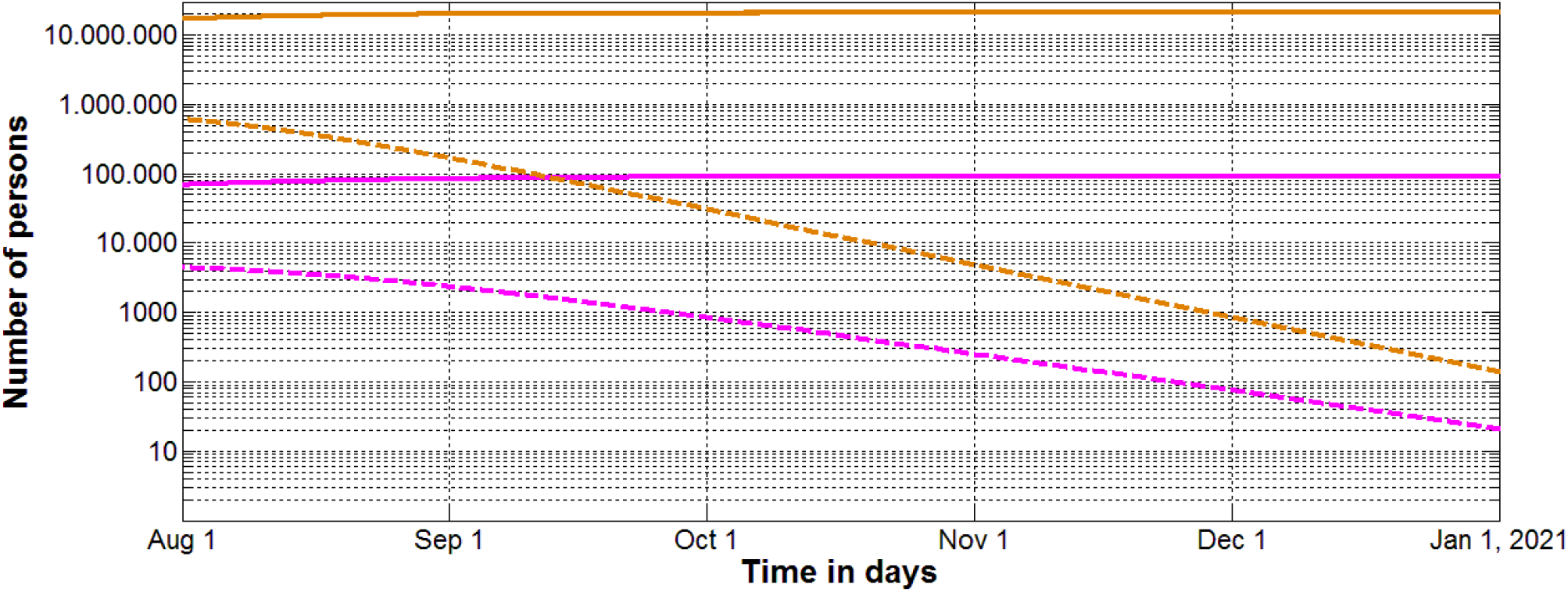
Long-term predictions for Ukraine (fourth wave) and the world (fifth wave). SIR curves (lines) and accumulated number of cases (markers) versus time. Magenta and brown colors correspond to Ukraine (Table 3) and the world (Table 5) respectively. Numbers of infected and spreading *I* (dashed lines) and victims (accumulated number of conformed cases) *V*=*I+R* (solid lines).

Very long duration of the pandemic requires correction of our behavior, we can not live as before it occurred. Decreased feelings of insecurity and non-compliance with social distancing may further increase the pandemic duration and the number of the coronavirus victims. Total closure of settlements or regions can be recommended only in the event of a sharp increase in the number of cases. There are many things that can be done without loss to the economy and our daily lives:

1. *Minimize the number of contacts and trips, not visit crowded places. Work and study remotely where possible*.
2. *Refrain from shaking hands and kisses during meetings. Use masks in transport and crowded areas*.
3. *If you* (*or others*) *have any suspicious symptoms, do your best to avoid the spread of the infection*.

It must be noted that the relatively small number of people who spread the coronavirus can cause a fairly long course of the epidemic. In particular, 20 such persons in Ukraine (estimation at the beginning of 2021, see Fig. 6) can cause new cases for another two and a half months. This explains the many new outbreaks in South Korea, Japan, China, Singapore and not all of them are caused by imported cases, [1, 21, 22]. To finally overcome the pandemic, it is very important to identify all the infected, and this is very difficult due to the large number of asymptomatic patients.

The dynamics of Covid-19 pandemic in different countries was simulated with the use of different mathematical models [23-37]. Nevertheless, in literature there are no long-time predictions for the final sizes and durations of the pandemic in Ukraine and in the world in order to compare with the presented results.

### Probability of meeting an infected person

As long as there are infected people in the population, there is a chance to meet one of them. The corresponding probability can be estimated with the use of simple formula:

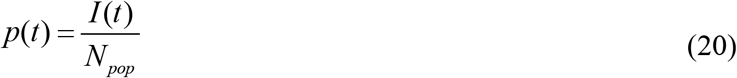

where *N*_*pop*_ is the number of persons in a country or in a region (volume of population). With the use of corresponding SIR curves it is possible to estimate *p*(*t*) for every moment of time. For example, on July 19, 2020 these probabilities can be estimated as 1.04·10^−4^ and 9.28·10^−5^ for Ukraine and the world respectively. It means that Ukraine was 12% more dangerous in comparison with the average situation in the world.

Formula (20) can be useful for deciding which countries’ citizens are welcome guests after the resumption of international passenger traffic. If *p*_*A*_ (*t*) > *p*_*B*_ (*t*) for countries A and B, citizens of A are not welcome guests in country B at the moment of time *t*. Since such decisions are very important, we will propose some simple method of estimating *p*(*t*) without calculations of SIR parameters for epidemic waves in a country. According to (13):

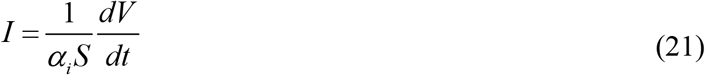

Since function *S*(*t*) changes not very sufficient during a wave (compare initial *S*_*i*_ =*N*_*i*_ *–I*_*i*_ *–R*_*i*_ and final *S*_*i*∞_ values of this function in Tables 2-5 and in corresponding Tables in [11, 15]), we can use the constant value *S*_*i*_ instead of *S*(*t*). To estimate *dV*/*dt*, it is possible to use the known values *V*_*j*_

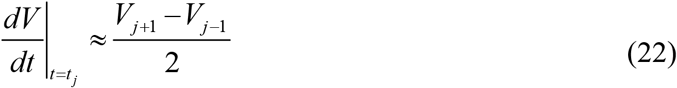

or formula (2) for smoothed number of cases. Finally, with the use of (21), equation (20) can be written as follows:

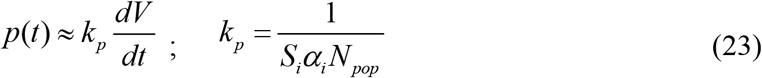

The results of calculations for different countries and regions are presented in Table 7. It can be seen that coefficient *k*_*p*_ changes not very fast (compare values on different days for Ukraine and the world). It means that values of this coefficient can be used during rather long period of time before new exact estimations of *I*(*t*) will be available. According to eq. (23), the probability of meeting an infected person mostly depends on the daily increase in the number of cases. The values *p* were estimated for July 19, 2020 and presented in Table 7.

**Table 7.**
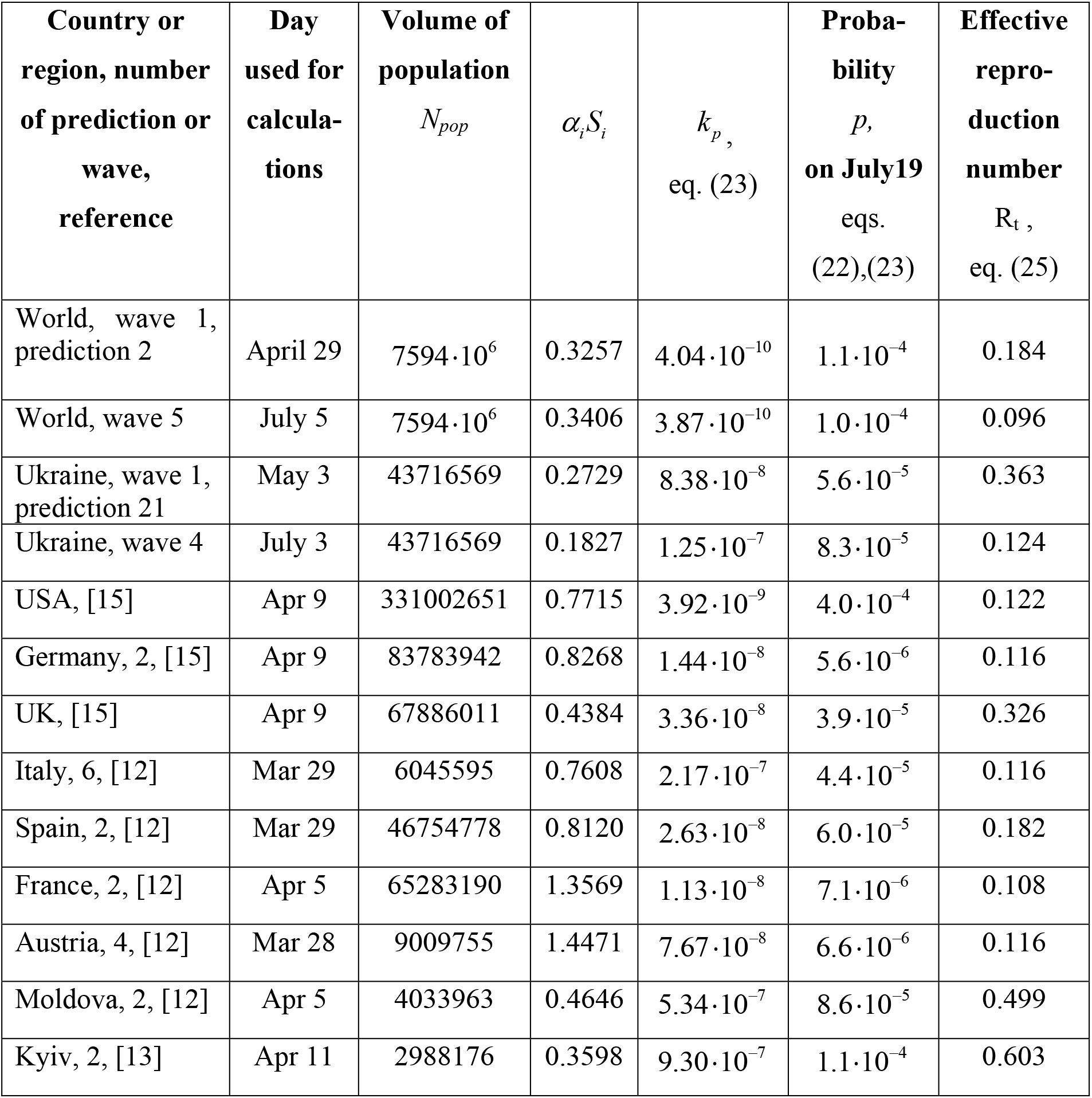
Epidemic characteristics for different countries and regions calculated for different days.

The *p* value for the fourth wave in Ukraine in 20% lower than the exact estimation 1.04·10^−4^ calculated before with the use of eq. (20). In the case of the world fifth wave, the difference between the exact estimation (20) and the approximate formula (23) is only 8%. It can be seen that the most infected country is USA. The average global figure is approximately 4 time less. The probabilities *p* in Kyiv, Moldova and Ukraine are around the world average value. Lower figures were calculated for Spain, Italy and UK. The safest counties are France, Austria and Germany.

### Estimations of reproduction numbers

Effective reproduction number *R*_*t*_(*t*) shows the average number of people infected by one person [38]. In terms of SIR model it can be estimated as:

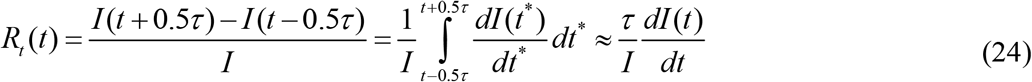

Where *τ* is the average time of spreading the infection. Taking into account (5), *τ* ≈ 1/ *ρ*_*I*_ constant value *S*_*i*_ =*N*_*i*_ *–I*_*i*_ *–R*_*i*_ instead of *S*(*t*), equation (24) can be rewritten as follows: and using

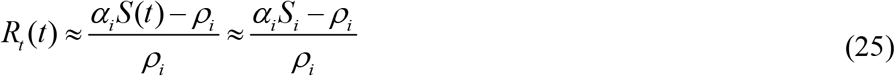

Equation (25) was used for calculations of the effective reproduction number for different regions and moments of time. The results are shown in the last column of Table 7.

It can be seen that France, Italy, Germany and Austria have the lowest values of *R*_*t*_ which are close to the world figure (wave 5). Not much higher values were calculated for USA and Ukraine (wave 4). Spain, UK, Moldova and Kyiv demonstrate the highest figures. The large difference in Ukrainian and Kyiv values can be explained by very different days of estimation (July 3 and April 11 respectively). Earlier periods of pandemic have higher values of *R*_*t*_. The same tendency demonstrates Table 7.

The *R*_*t*_ values corresponding to July 19, 2020, were estimated for Ukraine (wave 4, Table 3) and the world (wave 5, Table 5) with the use of exact dependences *I*(*t*) in formula (24) in order to compare with the results presented in Table 7. The values *R*_*t*_ = 0.0257 and *R*_*t*_ = 0.0059 for Ukraine and the world respectively demonstrate that the effective reproduction number can decrease very rapid over time.

The effective reproduction numbers of Covid-19 pandemic were estimated with the use of different mathematical models for different countries [37, 39, 40]. In particular, in [37] the *R*_*t*_ values were estimated as of May 10 for EU countries. Corresponding values for Austria, Spain, Germany, France and Italy vary from 0.45 to 0.74 and significantly exceed those shown in Table 7. Rapid changes in *R*_*t*_ values for the epidemic in China are reported in [39] (from 2.35 on January 16 to 1.05 on January 31).

The basic reproduction number *R*_*0*_ for the initial periods of an epidemic outbreak can be estimated with the use of equation, [38]:

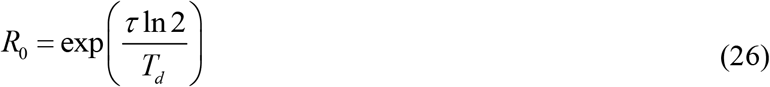

where *T*_*d*_ is the doubling time (the period during which the number of cases duplicates; the exponential growth is assumed). In the first half of March 2020, the increase in cases in the United States and around the world was close to exponential, [10]. Taking into account the values *T*_*d*_ = 2.31 and *T*_*d*_ =3.65 days for USA and the world respectively (calculated in [10]) and estimation *τ* ≈ 1/ *ρ*_1_, the values *R*_*0*_ =1.55 (USA) and *R*_*0*_ =1.99 (the world) can be obtained (we have used eq. (26) and values *ρ*_1_ = 0.6877, [15]; *ρ*_1_ = 0.275, Table 4).

The basic reproduction numbers of Covid-19 pandemic were estimated in [29, 41-44]. According to WHO calculations for China [41], values *R*_*0*_ vary from 2 to 2.5. During the initial period of an epidemic outbreak many infected persons are undetected. This fact diminish the accuracy of *R*_*0*_ estimations. For example, paper [44] reported the *R*_*0*_ =7.97 for Indonesia and two different values 1.54 and 6.22 for Singapore.

## Conclusions

In order to detect the Covid-19 pandemic waves, a simple method was proposed and used for Ukraine and the world. The SIR model and statistical approach to the parameter identification were modified and some reliable estimations of the epidemic waves are presented. In particular, the optimal values of the SIR model parameters were calculated for four pandemic waves in Ukraine and five waves in the world. E.g., the first wave dynamics allows estimating the real time of the outbreak. To calculate the final size and the duration of the epidemic, simulations of its next waves were used. The number of cases and the number of patients spreading the infection versus time were calculated.

The results demonstrate that the pandemic probably began in August 2019. If current trends continue, the end of the pandemic should be expected no earlier than in March 2021 both in Ukraine and in the world, the global number of cases will exceed 20 million. The probabilities of meeting a person spreading the infection and reproduction numbers were calculated for different countries and regions. The obtained information will be useful to regulate the quarantine activities, to predict the medical and economic consequences of the pandemic and to decide which countries’ citizens are welcome guests.

## Data Availability

Data are in text

## Acknowledgements

I would like to express my sincere thanks to Gerhard Demelmair, Anatolii Nikitin and Anatolii Podkur for their help in collecting and processing data.

